# Age-specific rate of severe and critical SARS-CoV-2 infections estimated with multi-country seroprevalence studies

**DOI:** 10.1101/2021.07.29.21261282

**Authors:** Daniel Herrera-Esposito, Gustavo de los Campos

## Abstract

Knowing the age-specific rates at which individuals infected with SARS-CoV-2 develop severe and critical disease is essential for designing public policy, for epidemic modeling, and for individual risk evaluation. In this study, we present the first estimates of these rates using multi-country serology studies, and data on hospital admissions and mortality from early to mid-2020. We integrated data from those sources using a Bayesian model that accounts for the high heterogeneity between data sources and for the uncertainty associated to the estimates reported from each data source. Our results show that the risk of severe and critical disease increases exponentially with age, but much less steeply than the risk of fatal illness. Importantly, the estimated rate of severe disease outcome in adolescents is between one and two orders of magnitude larger than the reported rate of vaccine side-effects, showing how these estimates are relevant for health policy. Finally, we validate our results by showing that they are in close agreement with the estimates obtained from an indirect method that uses reported infection fatality rates estimates and hospital mortality data.

## Introduction

The SARS-CoV-2 pandemic had impacts of historic proportion in both public health and society. Remarkably, there is considerable uncertainty regarding the full spectrum of health effects of SARS-CoV-2 infection. On the one hand, some of the effects of SARS-CoV-2 are relatively well understood, such as the infection fatality rate (IFR) and its dependence on age. Three different meta-analyses now exist that estimate the age-stratified IFR of SARS-CoV-2 using multi-country seroprevalence studies (Levin et al, O’Driscoll et al, Brazeau et al). These studies document an exponential increase of the IFR with age and show considerable agreement on their estimated IFRs by age-stratum. On the other hand, the rate of less extreme infection outcomes, and their dependence on age, remains uncertain despite being similarly important for public health. Examples of this are the rate of severe infections (Infection-severe rate, ISR), which we define as infections resulting in hospitalization or out-of-hospital death, and the rate of critical infections (Infection-critical rate, ICR), which we define as infections resulting in critical care admission or out-of-ICU death.

Knowing the age-specific ISR and ICR is essential for many reasons. One particularly important and pressing example where this information is required is in defining recommendations of vaccination of children and adolescents. Recent reports point to a link between mRNA vaccines and myocarditis (a relatively mild condition) in young people, even if at low rates (Bozkurt et al., 2021; Montgomery et al., 2021). While the CDC estimated the rate of vaccine-induced myocarditis to be approximately 1 in 80,000 second doses for individuals between 16 and 30 years old (Bozkurt et al., 2021), the FDA has estimated this rate at 1 in 5,000 vaccinated males between 16-17 years old, the highest risk group (FDA, n.d.), in line with the estimate of 1 in 6637 second doses for males between 16-19 years old reported in Israel (Mevorach et al., 2021). This, together with the low risk of death that children and adolescents face from COVID-19 (Brazeau et al., 2020; Levin et al., 2020; O’Driscoll et al., 2020), has raised questions about the cost-benefit balance of vaccinating young individuals (Vogel & Couzin-Frankel, 2021). But despite their relevance to this discussion, estimates of ISR and ICR for these ages using multi-country data are still missing from the literature (see estimates of the infection-hospitalization rate, for France in (Hozé et al., 2021; Lapidus et al., 2021), for Denmark in (Espenhain et al., 2021), for Indiana, USA, in (Menachemi et al., 2021), for Connecticut, USA, in (Mahajan, Caraballo, et al., 2021), for Qatar in (Seedat et al., 2021), and a model-based analysis with early non-serological pandemic data from (Verity et al., 2020).

To fill this gap, we present a meta-analysis of the age-stratified rates of severe and critical disease of SARS-CoV-2 across several locations, combining seroprevalence studies from early to mid 2020 with public data on the numbers of age-stratified hospitalizations, ICU admissions, and deaths.

## Results

We analyzed locations with seroprevalence studies that were either listed in the meta-analysis of Levin et al. (2020), to which we refer for further details, or the studies providing their own age-stratified rates of infection-hospitalization rate. We included in the analysis 15 locations with serosurveys (11 using representative samples and 4 using convenience samples), and 2 locations with comprehensive testing and contact tracing, which we corrected for under-ascertainment using seroprevalence data from Iceland, following Levin et al. (2020). Together, these locations represent 5% of the world’s population.

Also, considering that out-of-hospital and out-of-ICU deaths are common among the elderly, we added the out-of-hospital and out-of-ICU deaths to the reported hospitalizations and ICU admissions, to obtain the severe and critical cases respectively. For some locations this information was available; for the other locations we estimated the out-of-hospital or out-of-ICU deaths using data on the total deaths and hospital and ICU mortality estimates.

The estimated probability of severe, critical, and fatal disease outcomes (ISR, ICR, and IFR respectively) are shown for each age and location as colored points in **Figure 1**, using a log-transformed vertical axis. As expected from the reports in previous analyses of IFR (Brazeau et al., 2020; Levin et al., 2020; O’Driscoll et al., 2020) and from reports on infection-hospitalization ratios (Hozé et al., 2021), the three outcome ratios show an approximately exponential increase in risk with respect to age, which becomes a homogeneous linear effect on the log-scale. We used this observation to fit models that allow us to estimate those ratios for all ages and locations together, improving the accuracy of estimates at young ages where these adverse outcomes are less frequent (similar to (Levin et al., 2020)). Specifically, for each of the infection rates (ISR, ICR, and IFR) we fitted Bayesian logistic regression models with a linear age effect on the logit scale (this effect becomes non-linear in the risk scale). We used logistic regression because it is a commonly used model for disease outcomes that approximate very well the log-linear rate-age patterns observed in our study. The random-effects logistic regression model had a linear component (intercept and age-slope) shared across locations, plus location-specific random effects on both the slope and the intercept of the regression. The location random-effects, together with a consideration of the uncertainty of seroprevalence estimates, and the variability associated with the random nature of the observed number of deaths, allowed us to rigorously combine the highly heterogeneous data sources from across locations.

**Figure 1.**
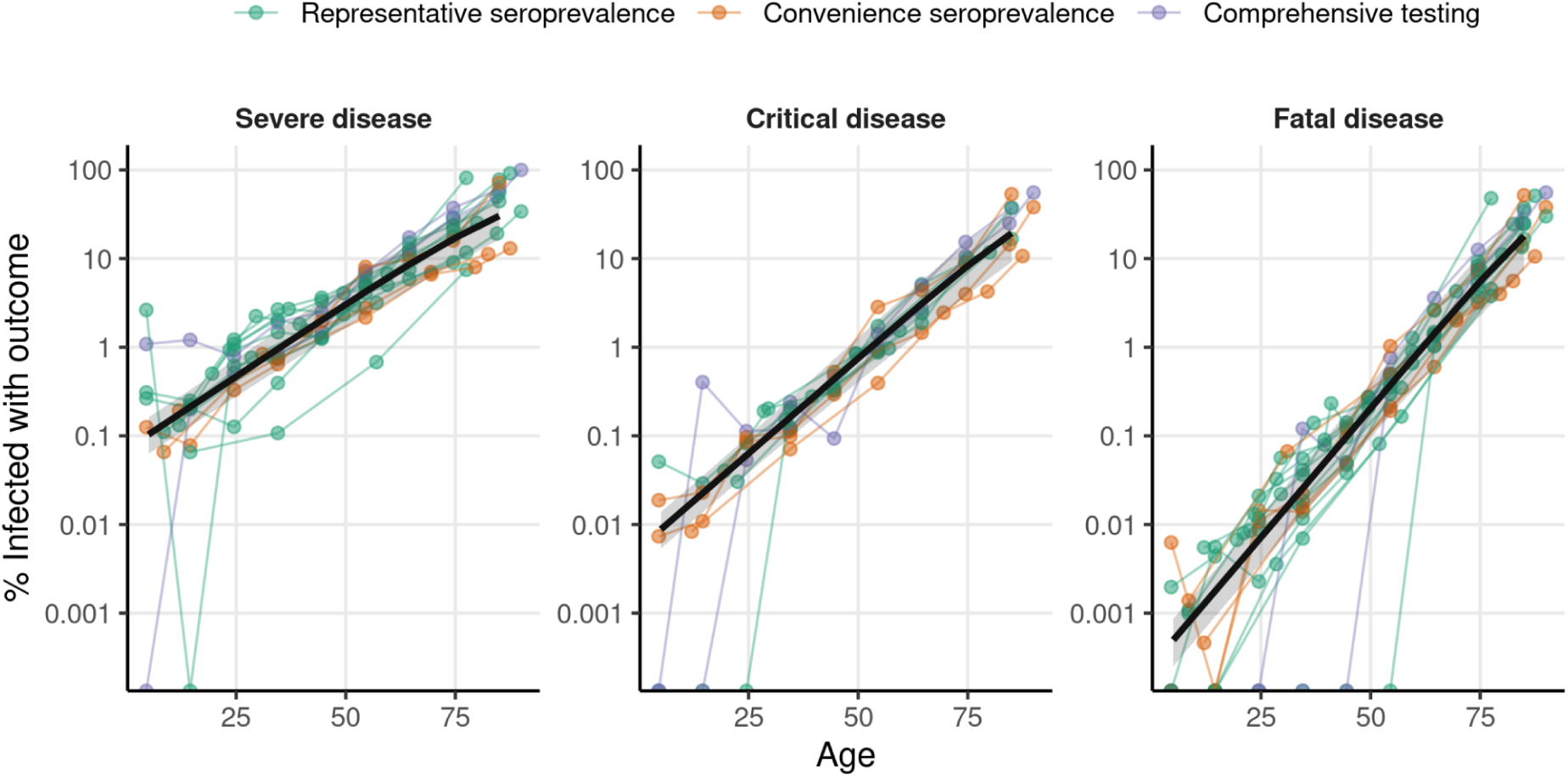
Rates of severe and critical SARS-Cov-2 outcomes (ISR and ICR, respectively) and death rates (IFR) estimated with seroprevalence data from 2020. The colored points show the proportion of individuals infected with SARS-CoV-2 that develop severe disease (left), critical disease (center), or fatal disease (right) (in logarithmic scale) for each location and age-stratum used in our analysis. Color indicates whether the number of infections were obtained from a representative serosurvey, a convenience serosurvey, or from comprehensive testing corrected for under-ascertainment. Data points coming from a given location are joined by colored lines. The black line shows the outcome rate estimated using a hierarchical Bayesian logistic regression model, and the shaded regions show the 95% credibility intervals. We used 105 data points from 16 locations for the estimation of ISR, 78 data points from 11 locations for ICR, and 119 data points from 17 locations for IFR.

The logistic regression models fitted the data well for the three outcome rates (**Figure 1**, although see the discussion of supplementary section **S1** about a possible deviation from the trend for the youngest ages), and the patterns observed were similar across locations. Importantly, the slope of IFR with respect to age (0.133, 95% credibility interval: [0.123-0.143]) was higher than the slope of ICR (0.099, [0.089-0.108]) and of ISR (0.076, [0.067-0.083]), indicating that the risks of severe and critical disease are more evenly distributed across ages than the risk of death. The models also had different intercepts, of −12.9 [−13.6, −12.3] for IFR, −9.9 [−10.3, −9.4] for ICR and −7.3 [−7.7, −6.8] for ISR, reflecting the difference of 1 order of magnitude between ICR and IFR for the youngest ages, and of 2 orders of magnitude between ISR and IFR. As an example, according to our estimates, people in the 20-25 years old range are on average 779 [467-1223] times less likely to die from COVID-19 than 70-75 year old people, 133 [83-202] times less likely to develop critical disease, and 38 [27-51] times less likely to develop severe disease. Predicted risk levels by age (and the corresponding 95% credibility intervals) are shown in **Table 1** (see **Table S1** for finer age stratification, and **Table S2** for the estimated model parameters). Also, we verified that our estimates are robust to the correction for out-of-hospital and out-of-ICU deaths (**Figures S1,S2**), the ages used to fit the model (**Figure S3**), the method of estimating SARS-CoV-2 infections (**Figure S4**), the date of outcome data collection (**Figures S5,S6**), and the delay between the epidemic wave and the seroprevalence study (**Figure S7**, to control for seroreversion). More details about these comprehensive robustness analyses are provided in the Supplementary Materials.

**Table 1.** Estimated age-specific frequencies of severe disease (ISR), critical disease (ICR), and fatal disease (IFR) among infected individuals. The estimates are obtained from the fits to the serology data from 2020 shown in **Figure 1**. Numbers in the parenthesis indicate 95% credibility intervals of the estimates, obtained by taking the 2.5% and 97.5% quantiles of the posterior probability of the bayesian fit. (*) Estimates for the youngest ages may be underestimated by the assumption of a logistic relation between age and severity, see supplementary section **S1** for further discussion and complementary estimates.

| Age | ISR % (CrI) | ICR % (CrI) | IFR % (CrI) |
| --- | --- | --- | --- |
| 0-9 | 0.103 (0.063-0.162) (*) | 0.0088 (0.0053-0.0139) (*) | 0.00050 (0.00025-0.00087) (*) |
| 10-19 | 0.22 (0.13-0.35) | 0.024 (0.014-0.037) | 0.0019 (0.0010-0.0033) |
| 20-29 | 0.47 (0.28-0.74) | 0.063 (0.038-0.10) | 0.0072 (0.0037-0.0126) |
| 30-39 | 0.99 (0.57-1.61) | 0.17 (0.10-0.28) | 0.027 (0.014-0.049) |
| 40-49 | 2.1 (1.2-3.5) | 0.46 (0.26-0.77) | 0.10 (0.05-0.19) |
| 50-59 | 4.4 (2.4-7.4) | 1.2 (0.6-2.1) | 0.40 (0.18-0.77) |
| 60-69 | 8.9 (4.6-15.2) | 3.3 (1.6-5.9) | 1.5 (0.6-3.0) |
| 70-79 | 17.1 (8.9-28.8) | 8.3 (3.9-15.5) | 5.5 (2.3-11.3) |
| 80+ | 30.3 (16.4-47.7) | 19.4 (9.2-34.7) | 18 (7.5-34.3) |

As a reference, when compared to the 1/5000 rate for mRNA-vaccine-induced myocarditis in 16-17 year-old males (FDA, n.d.), our estimate of the rate of severe disease for this age range is 12 [7-19] times larger, the rate of critical disease is 1.4 [0.8-2.2] times larger, while the rate of fatal disease is 0.12 [0.07-0.20] times the rate of vaccine induced myocarditis. Using the alternative rate of myocarditis of 1/80,000 estimated by the CDC (Bozkurt et al., 2021) for the 16-30 age group, the rate of severe disease is 322 [193-506] times larger, the rate of critical disease 42 [25-66] times larger, and the risk of death is 4.4 [2.3-7.7] times larger. We emphasize that such comparisons do not constitute a proper risk-benefit analysis, since such an analysis ought to consider the risk of infection for both vaccinated and unvaccinated individuals, as well as the probability of becoming infected during the protective period of the vaccine, among other factors. However, the difference of one or two orders of magnitude (depending on the rate of vaccine-induced myocarditis used) that we observe between ISR and the rate of myocarditis highlights the importance of taking into account the rates of severe disease for these age ranges, particularly considering that the reported cases of vaccine-induced myocarditis have been mostly mild (Mevorach et al., 2021; Montgomery et al., 2021; Vogel & Couzin-Frankel, 2021). These estimates thus provide an important input to such more elaborate risk-benefit analyses.

Next, we validated our estimates by estimating the ISR and ICR of SARS-CoV-2 indirectly using a novel ratio-of-ratios approach. We start from the age-specific IFR reported in the three different meta-analyses (Brazeau et al., 2020; Levin et al., 2020; O’Driscoll et al., 2020), which were not used in the analysis of **Figure 1**. Because the age-specific IFR is the expected ratio between deaths and infections for a given age, if we know the expected ratio between deaths and severe infections for this age (the severe fatality rate, SFR), we can then obtain the age-specific ISR by taking (the age-specific) ratio IFRs/SFRs. To approximate the SFR, we used data on hospital mortality for COVID-19 patients, which is the ratio between in-hospital deaths and hospitalizations. This approximation is expected to hold well for all but the oldest age bins (see **Supplementary Section S2, Figure S2**). We fitted a Bayesian logistic regression model (like the one used to generate the results in **Figure 1**) to hospital mortality data of COVID-19 patients (**Figure 2A**, data sources listed in **Table M3**). Then, we took the ratio between the age-specific IFRs and the estimated age-specific hospital mortalities to obtain ISR estimates. We then applied the same procedure using the mortality of COVID-19 patients in critical care, to obtain estimates of the ICR. The values of the parameters obtained by fitting the model to hospital and ICU mortality are shown in **Table S3**, and the age-specific estimates are shown in **Table S4**.

**Figure 2.**
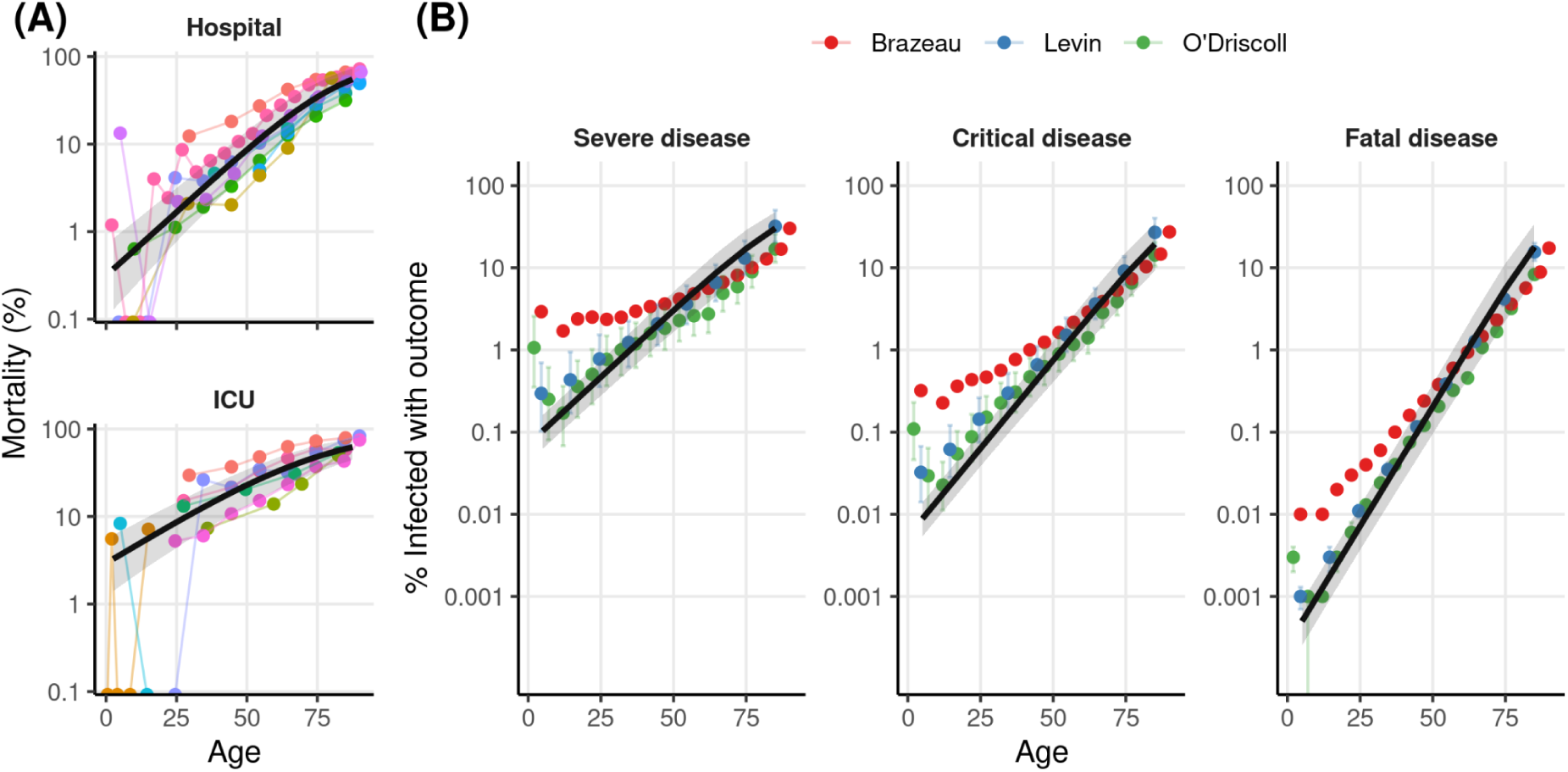
Rates of severe (ISR) and critical (ICR) obtained with an indirect method based on a ratio of ratios. **A)** The colored points represent the reported mortality rates of hospitalized (top) and ICU SARS-Cov-2 patients, each study is reported in a different color. The black line shows the estimated outcome rates for each age obtained from our hierarchical Bayesian logistic regression, and the shaded regions show the 95% credibility intervals. 68 data points from 8 reports were used for hospital mortality, and 43 data points from 8 reports were included for ICU mortality. **B)** The colored points show the estimated rates of severe (left) and critical (center) disease, obtained by dividing the age-stratified IFRs of the three relevant meta-analyses (Brazeau et al., 2020; Levin et al., 2020; O’Driscoll et al., 2020) by the corresponding values obtained in **A)**. The points show the mean values of the posterior distribution, and bars show 95% credibility intervals (we omit these for Brazeau et al. since the credibility intervals around the mean estimates are not reported). The rightmost plot shows the IFRs reported by each of the studies. The black line and shaded region in each panel show the meta-analysis estimates we obtained with the direct method (**Figure 1**).

We note two relevant details from this method. First, we used the same hospital and ICU mortality data for this analysis and for correcting for out-of-hospital and out-of-ICU deaths in **Figure 1**. Although this means that the two analyses share some data in common, the results of the regression of **Figure 1** are similar when performed on the uncorrected data (**Figures S1,S2**), supporting the use of this validation method. Second, although it is possible to correct for out-of-hospital and out-of-ICU deaths, instead of only approximating the SFR as the hospital mortality, the effect is expected to be small for most ages, and such a correction would involve using some of the same data as in **Figure 1**. Thus, to keep the two estimation methods as independent as possible for the purpose of validation, we chose not to correct for out-of-hospital and out-of-ICU deaths.

The estimates from the indirect method are shown by the colored points in **Figure 2B**. To aid comparison, we show in black the lines obtained from the fit to serology data of **Figure 1**. Firstly, we see that our IFR estimates and the IFR estimates from the three meta-analyses are very similar (**Figure 2B, right**). Second, we note that the estimates of ISR and ICR obtained from seroprevalence and disease outcome data with the direct method (**Figure 1**) are in close agreement with the estimates obtained with the indirect method (with the largest differences being with Brazeau et al. (2020) for the younger ages; however, this study also reports estimates different than those reported in the other two studies).

## Discussion

In this work, we present the first meta-analysis of serology studies for estimating age-specific ISR and ICR of SARS-CoV-2 during early to mid 2020. Our estimates show that, as with other SARS-Cov-2 outcomes, the ISR and ICR increase exponentially with age; however, the rate of increase in the risk of severe and critical disease outcomes with age is less marked than the rate of increase in lethality due to SARS-Cov-2 infection. This result is in agreement with previous studies reporting smaller effects of age on the hospitalization-infection ratio (Blackburn et al., 2021; Espenhain et al., 2021; Lapidus et al., 2021; Mahajan, Caraballo, et al., 2021; Menachemi et al., 2021; Seedat et al., 2021; Verity et al., 2020), and on the ICU-infection ratio (Menachemi et al., 2021; Seedat et al., 2021) as compared to IFR. However, despite the general agreement between previous serology studies (which show a deviation from the model-based estimates of Verity et al. 2020), they show considerable variability, probably due to the uncertainty in serology estimates, differences in local reporting protocols, and geographical variability in the impacts of COVID-19–the Bayesian model used in this study allowed us to produce estimates that borrow information across locations while accounting for heterogeneity in disease outcomes across locations.

Furthermore, we validated our estimates through several robustness analyses (see **Supplementary materials**) as well as using a novel ratio-of-ratios method to validate our estimates using different data sources (i.e. hospital and ICU mortality reports). Our simple Bayesian regression analysis that takes into account several sources of uncertainty and this novel ratio-of-ratio methods constitute two complementary methods that may aid future work on estimating these parameters under the changing nature of the pandemic.

The results presented in this study are an essential input for the cost-benefit analyses that many countries need to conduct when making policy decisions. For example, our estimates are an essential input for assessing the impacts of recommendations regarding children and adolescent vaccination–a problem of great public health relevance with potential impacts not only children and adolescents’ health but also on the future trajectory of the pandemic. Also, besides their use for determining official public-health recommendations, our results can also be used to develop a more complete and accurate public communication campaign of COVID-19 health risks for young and middle-aged individuals, who are the most hesitant about COVID-19 vaccination (Murphy et al., 2021).

Our results are also highly relevant for other aspects of COVID-19 modeling, such as estimating the number of unreported infections from hospital and ICU data, allowing to better estimate the present levels of natural immunity (Hozé et al., 2021; Irons & Raftery, 2021; Russell et al., 2020); prediction of the effects of public-health policies implemented along the pandemic (Davies, Barnard, et al., 2021); evaluating policy decisions such as vaccine allocation (Matrajt et al., 2020); the prediction of health outcomes in countries with high or low vaccination rates that account for the age-distribution of each country (Davies et al., 2020; Sandmann et al., 2021).

Finally, we note that the dynamic nature of the COVID-19 pandemic makes any estimates of outcome’s rate transient, since those rates are expected to change in space in time as new variants emerge and social behavior and medical practices change. As such, generalization of our estimates across time and space requires caution. For example, a drop of around 50% in hospital mortality between the first and second waves has been reported in developed countries, (Anesi et al., 2021; Asch et al., 2021; Aznar-Gimeno et al., 2021; Navaratnam et al., 2021), and ICU mortality was reported to drop 12% in the UK (Doidge et al., 2021). How the situation has continued to change is unclear, although a meta-analysis of ICU mortality suggests that improvements may have plateaued between June and September 2020 (Armstrong et al., 2021). Also, there is considerable variability of the improvements in mortality across hospitals in the US (Kadri et al., 2021), outlining the spatial heterogeneity in how these rates change over time. The spatial heterogeneity is likely also accentuated by differences in reporting or admission protocols (eg. due to healthcare demand and testing capabilities), both across locations and across time, which may also affect any individual estimate of disease severity, as well as our meta-analysis.

Furthermore, besides the effects of improvements in treatment, the emergence of variants of concern may have conversely increased the rates of adverse disease outcomes. Different studies associate the emergence of variants of concern to increased hospital mortality (Freitas et al., 2021; Jassat et al., 2021; Khedar et al., 2021), as well as to increases in severe, critical and fatal disease among reported cases (Challen et al., 2021; Davies, Jarvis, et al., 2021; Fisman & Tuite, 2021a, 2021b; Grint et al., 2021; Ong et al., 2021; Tuite et al., 2021).

Thus, although it is to be expected that the rates estimated in our work have changed during the pandemic, the extent of this change given by the combination of the many relevant factors is uncertain. Furthermore, estimating such changes is challenging, given the large heterogeneity of situations across countries, as well as other technical difficulties, such as accounting for the effects of growing population immunity. However, the estimates presented here are the most up to date estimates derived from the best available data from multiple locations worldwide. Therefore, the results reported here constitute an important reference of the health impacts of COVID-19 during 2020, as well as an important baseline over which to build more updated estimates, by combining them with estimates of the relative change in risk across locations and time.

## Materials and Methods

The data sets included in this study come from locations where age-stratified seroprevalence studies have been performed (see M1), plus locations with age-stratified prevalence coming from exhaustive contract tracing (see M2). For each of these locations, we searched for age-stratified data on Hospitalizations and ICU admissions (see M3). We used these two sources of data to estimate the age-stratified rates of severe (ISR) and critical (ICR) SARS-CoV-2 infections for each of the locations. We used this data to fit Bayesian random-effects logistic regression models for each of the outcomes (see M4). We also searched for studies reporting age-stratified mortality for COVID-19 patients admitted to the hospital or to the ICU (see M5). We used this data to fit a Bayesian random-effects logistic regression model to obtain the age-specific hospital and ICU mortality for COVID-19. This regression was then combined with estimates of age-specific IFR extracted from the literature, to estimate the ISR and ICR through a ratio-of-ratios method (see M6). The regression of hospital and ICU mortality was also used to correct the hospital and ICU data described in M3 for out-of-hospital and out-of-ICU deaths (see M7). All data and code are available online (see M8).

### M1) Data from seroprevalence studies

We used a curated list of seroprevalence studies released prior to 18 September 2020 that is presented in (Levin et al., 2020)–a systematic review and a meta-analysis. The list is restricted to developed countries; we refer the reader to (Levin et al., 2020) for an exhaustive list of other existing studies, and the criteria used for excluding seroprevalence studies from their final analysis.

We then searched for age-stratified hospitalization and ICU data to match the representative seroprevalence studies listed in Appendix Tables I.1 and I.3, and the convenience seroprevalence studies listed in Appendix Table I.2 from Levin et al. (2020). From the 11 representative seroprevalence studies included in those lists, we were able to find age-stratified hospitalization or ICU data for 7 locations (England; France; Ireland; Netherlands; Spain; Atlanta, USA; Geneva, Switzerland) and we failed to find such data for the 4 remaining locations (Italy; Portugal; Indiana, USA; Salt Lake City, USA). From the 4 convenience seroprevalence studies listed, we were able to find hospitalization or ICU data for all three locations (Ontario, Canada; Sweden; Belgium; New York, USA).

In addition to the seroprevalence studies used in (Levin et al., 2020), we included the seroprevalence study carried out in Iceland up to April 4 2020, which reports the results of a representative sample of the population (Gudbjartsson et al., 2020), and the representative seroprevalence studies from Indiana, Connecticut and Denmark, which were used to estimate the ISRs in the existing literature (Espenhain et al., 2021; Mahajan, Caraballo, et al., 2021; Menachemi et al., 2021).

**Table M1.** List of sources for the seroprevalence data of each location. The final date of the data collection period is shown in the center column.

| Location | End date | Source |
| --- | --- | --- |
| England (< 17 years) | 03/07/2020 | (Waterfield et al., 2021) |
| England | 13/07/2020 | (Ward et al., 2021) |
| Ile-de-France | 23/06/2020 | (Carrat et al., 2020) |
| Ireland | 16/07/2020 | (Ireland Health Service Executive, 2020) |
| Netherlands | 11/05/2020 | (Vos et al., 2020) |
| Spain | 11/05/2020 | (Pollán et al., 2020) |
| Atlanta, USA | 03/05/2020 | (Biggs, 2020) |
| New York City, USA | 28/04/2020 | (Rosenberg et al., 2020) |
| Ontario, Canda | 31/07/2020 | (Ontario Public Health, 2020) |
| Sweden | 24/05/2020 | (Sweden Public Health Authority, 2020) |
| Iceland | 04/04/2020 | (Gudbjartsson et al., 2020) |
| Geneva, Switzerland | 06/05/2020 | (Perez-Saez et al., 2020) |
| Belgium | 26/04/2020 | (Herzog et al., 2020) |
| Connecticut, USA | 29/07/2020 | (Mahajan, Srinivasan, et al., 2021) |
| Indiana, USA | 29/04/2020 | (Menachemi et al., 2021) |
| Denmark | 16/12/2020 | (Espenhain et al., 2021) |

Furthermore, in three cases we also changed the use of some seroprevalence studies with respect to Levin et al., (2020), to match them to the available hospitalization and ICU data. The first case is the seroprevalence study from France, which offers data by region. We were only able to find the age-stratified hospitalization data for the region of Île-de-France; therefore, we only used seroprevalence data from this region. The second case is the New York seroprevalence study, where we could only find hospitalization data for New York City but not for New York State; thus, we only used the seroprevalence for New York City. For Ontario, Canada, we could only find age-stratified hospitalization and ICU data up to July 31 2020, and so we used the seroprevalence report for this date, which is different from the report date used by (Levin et al., 2020). **Table M1** summarizes the final list of seroprevalence studies included in our analysis.

#### Matching age-bins

The age-bins reported by each of the studies did not always match the age-bins in the corresponding hospitalization and ICU reports. Therefore, in some cases we extrapolated or interpolated the seroprevalence estimates obtained for a given age-bin into a different age-bin. For example, for New York City, seroprevalence was reported for the 18-34 years old age range, but hospitalization data was reported for the 18-44 year old age range. Therefore, to make use of this hospitalization data, we assumed that the proportion of seropositive individuals in the 18-44 years old range is the same as the proportion for the 18-34 year old age range. All such decisions were contrasted with other available data, and agreed upon by the two authors. Furthermore, these assumptions are all documented in the publicly available analysis code.

#### Correcting for test characteristics

The positive rate of a test depends on disease prevalence and on the test characteristics. Most of the seroprevalence estimates used were already corrected for test characteristics. For the results that were not corrected for test characteristics, we used the Gladen-Rogan formula (ROGAN & GLADEN, 1978) to adjust the estimates as follows:

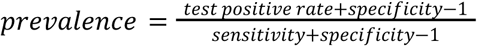

### M2) Countries with comprehensive tracing included in the analysis

Following (Levin et al., 2020), we also included in our analysis two countries (Republic of Korea and New Zealand) with comprehensive tracing programs where the number of infections detected through testing are thought to approximate the total number of infections accurately. As in the original Levin et al. study, we corrected the prevalence estimates for these countries using the age-specific ratio between the number of infections estimated through seroprevalence and the number of positive tests in Iceland (Gudbjartsson et al., 2020).

### M3) Hospitalizations, ICU admissions, and deaths data

We obtained the age-stratified hospitalizations, ICU, and death data in relevant government websites of the locations, using google search, and looking for relevant region-wide studies. We selected the data reports that were closest to the end of the serosurvey date. The list of data sources and the end dates for their cumulative outcome numbers are shown in **Table M2**.

**Table M2.**
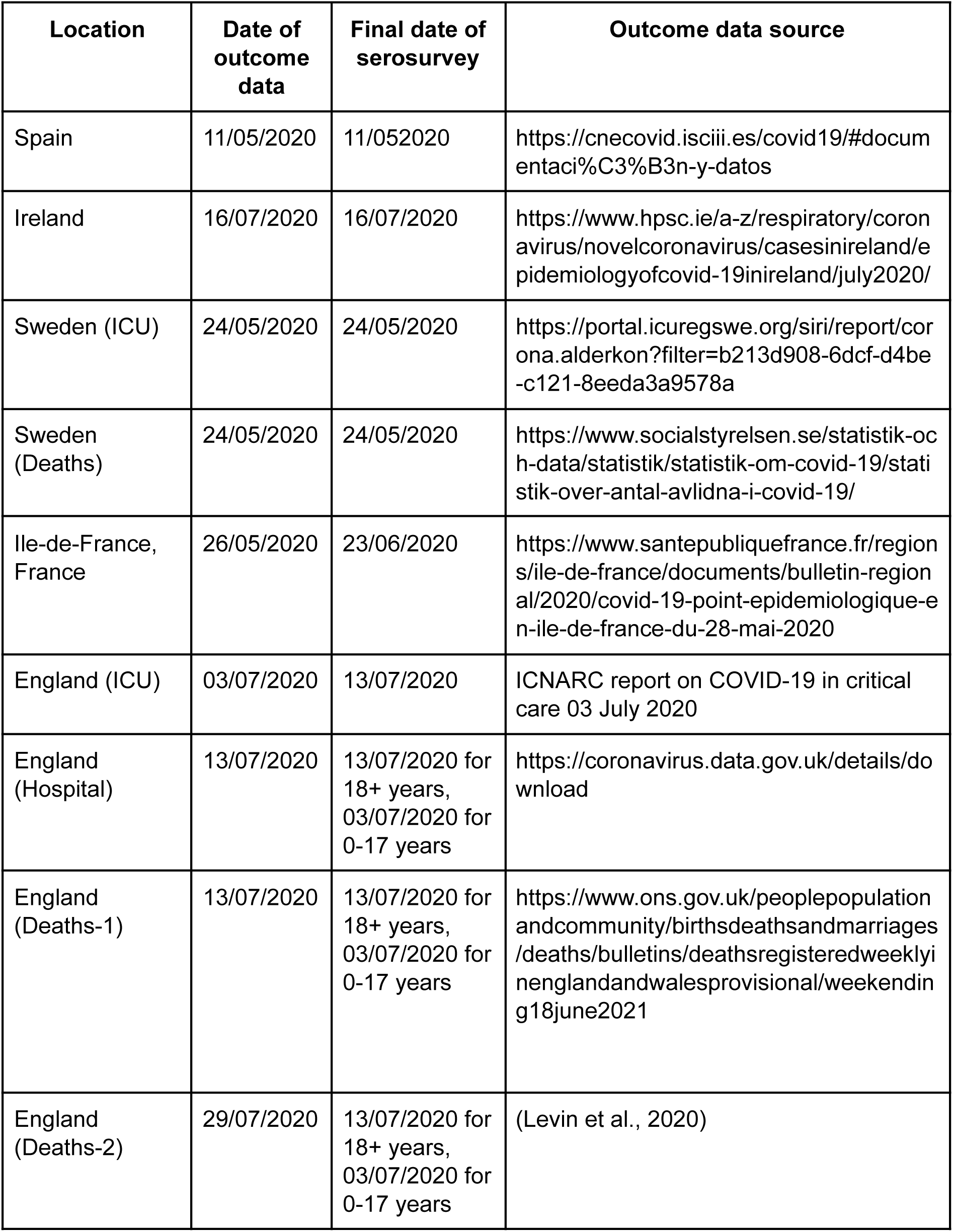
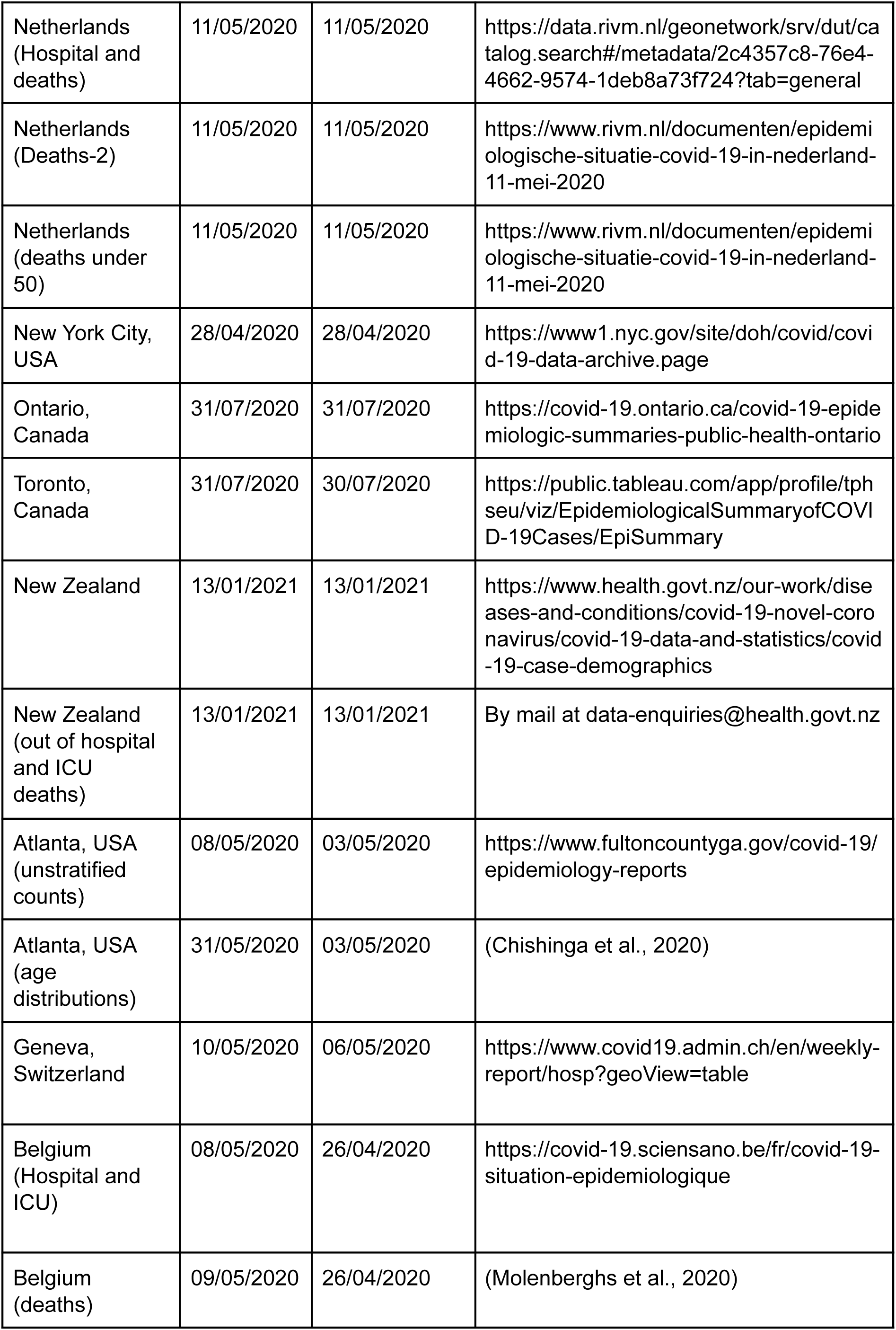
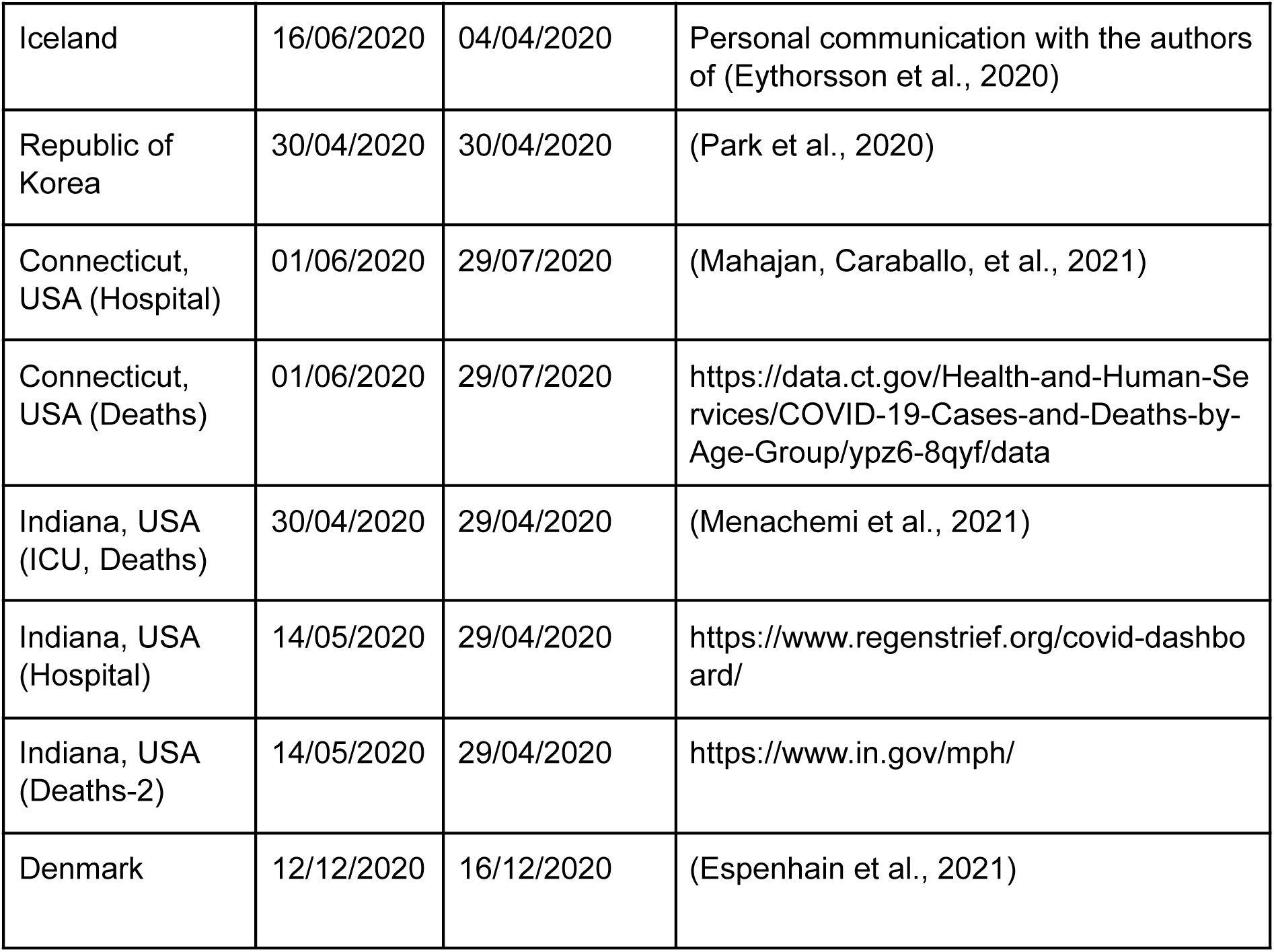
List of sources for the hospitalization, ICU, and death data for each location. The date up to which the cumulative numbers for these outcomes were reported are shown in the second column.

Also, as described above for the serology data, in some cases we interpolated or extrapolated some data for these disease outcomes, or we combined different data sources with incomplete data (e.g., age-distribution of an outcome from one source, with the total count of the outcome from another source) to obtain the data for these outcomes with the appropriate age bins. For example, for Belgium, we were only able to find the unstratified number of cumulative ICU admissions at the desired date of May 8^th^, 2020, but we were able to find the age distribution for cumulative ICU admissions up to June 14^th^, 2020. Therefore, we distributed the cumulative ICU admissions of May 8^th^ across age strata, following the distribution from June 14^th^. As mentioned above, all these decisions were agreed upon by the authors, and they are thoroughly documented in the publicly available analysis code.

### M4) Estimation of outcome probabilities with serology and outcome data

We fitted Bayesian logistic regression models to the serology and outcome data. We describe the model for severe SARS-CoV-2 outcome; the same model was fitted to severe, critical, and fatal disease outcomes.

Let {*y*_*la*_, *x*_*la*_} represent the number of severe SARS-Cov-2 infections (*y*_*la*_) experienced among *x*_*la*_ individuals infected with SARS-CoV-2, at the location *l* (*l* = 1, …, L), for the age stratum *a* (*a* = 1, …, A_*l*_) of the *l*^*th*^ location. The ISR for this location-stratum is defined as 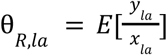.

#### Bayesian likelihood

The probability of *y*_*la*_ given θ_*R,la*_ and the number of infections (*x*_*la*_), is given by the Binomial likelihood 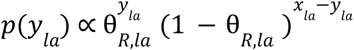. Assuming conditional independence across locations and strata, and taking 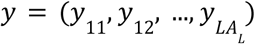, and 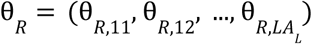 we have 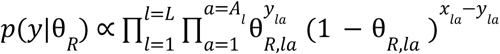

#### Modeling the number of SARS-Cov-2 cases from seroprevalence data

The selected seroprevalence studies provide age-stratified estimates (and SE) of disease prevalence. Rather than assuming that prevalence was known with complete certainty, we used the reported point estimates and SE to specify a Beta prior for prevalence for each location. Specifically, we used the reported prevalence and its SE to estimate (through first and second moments matching) the shape parameters of the Beta distribution used for each location. Then, prevalence was modeled as θ_P,*la*_ ∼*Beta*(α_1,*la*_, α_2,*la*_), where α_1,*la*_ and α_2,*la*_ are location-age-stratum specific shape parameters. Then, the number of cases was defined as *x*_*la*_ = *N*_*la*_ × θ_P,*la*_, where *N*_*la*_ is the size of the population at location *l* and age stratum *a*.

#### Modeling severity rates using random-effects logistic regression

Infection-severity rates were modeled using a logit of the form

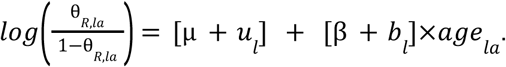

Therefore, 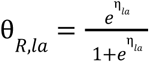 where *η*_*la*_ *=* [μ + *u*_*l*_] + [β + *b*_*l*_]×*age*_*la*_.

Above, µ and β are the average intercept and slopes across locations, and *u*_ij_ and *b*_ij_ are location-specific random effects on the intercept and the slope, respectively, with prior distribution 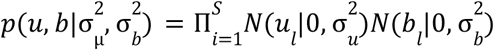. The shared intercept (µ) and regression coefficient (β) were assigned flat priors, and the standard deviations for the random effects, σ_u_ and σ_b_ were assigned gamma priors with shape and rate parameters equal to 4.

For each age stratum *a* at location *l*, the value of *age*_*la*_ used for fitting corresponded to the median age of the stratum. For age strata with an open upper bound (e.g. 70+ age), we used 90 years as the upper bound of the stratum.

The posterior distribution of the model described above does not have a closed form; therefore, we used Monte Carlo Markov Chain (MCMC) methods to generate samples from the posterior distribution for all the model unknowns 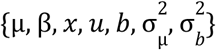. We used 4 chains with 2500 iterations each. A script that implements the above model in Stan (Carpenter et al., 2017) is available in the online code.

#### Prediction of outcome rates

We used the samples of the posterior distribution to generate posterior samples for the infection severity rates for specific ages using the inverse-logit function: 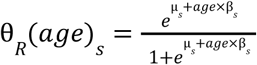, where *s* is an index for the sample from the posterior distribution. We then used these samples to estimate the posterior means and posterior credibility regions reported in Figures 1 and 2. We report the severity rates for age intervals by estimating the rate of the mean age of the interval.

The predicted outcome rates obtained from the model fit are shown in **Table S1**, and the mean and credible intervals for the main model parameters are shown in **Table S2**.

### M5) Hospital and ICU mortality data

Our robustness analysis was based on an indirect estimator (a ratio-of-ratios) of ISR and ICR. To derive this estimator, we used mortality data from hospitalized and ICU SARS-Cov-2 patients. We searched in the literature for reports on age-stratified mortality of patients admitted to the hospital or the ICU with a COVID-19 diagnosis. We also used the data sources from **Table M2** that provided mortality numbers for hospitalized or ICU patients. We identified 8 ICU mortality reports and 8 mortality hospital reports with age-stratified data, which were either published studies in the literature or public reports from official organisms. The reports used are listed in **Table M3**.

**Table M3.** List of sources for mortality among COVID-19 patients in the hospital or in critical care. The end date of each study is shown in the third column.

| Location | Patient type | Date | Source |
| --- | --- | --- | --- |
| England | ICU | 03/07/2020 | ICNARC report on COVID-19 in critical care 03 July 2020 |
| New York City, USA | ICU | 28/04/2020 | (Cummings et al., 2020) |
| France, Belgium, Switzerland | ICU | 04/05/2020 | (COVID-ICU Group on behalf of the REVA Network and the COVID-ICU Investigators, 2021) |
| Sweden | ICU | 01/09/2020 | https://portal.icuregswe.org/siri/report/corona.alderkon?filter=b213d908-6dcf-d4be-c121-8eeda3a9578a |
| Brazil | ICU | 15/08/2020 | (Ranzani et al., 2021) |
| Florida, USA | ICU | 18/05/2020 | (Oliveira et al., 2021) |
| Intercontinental | ICU | 23/04/2020 | (González-Dambrauskas et al., 2020) |
| Brazil | ICU | 31/05/2020 | (Prata-Barbosa et al., 2020) |
| New York City, USA | Hospital | 04/04/2020 | (Richardson et al., 2020) |
| Germany | Hospital | 19/04/2020 | (Karagiannidis et al., 2020) |
| France | Hospital | 13/05/2020 | (Salje et al., 2020) |
| United Kingdom | Hospital | 19/04/2020 | (Docherty et al., 2020) |
| Spain | Hospital | 17/04/2020 | (Berenguer et al., 2020) |
| Chile | Hospital | 04/06/2020 | (Maquilon et al., 2020) |
| Brazil | Hospital | 15/08/2020 | (Ranzani et al., 2021) |
| Netherlands | Hospital | 11/05/2020 | https://data.rivm.nl/geonetwork/srv/dut/catalog.search#/metadata/2c4357c8-76e4-4662-9574-1deb8a73f724?tab=general |

### M6) Indirect estimation of ISR and ICR using IFR and hospital mortality data

To validate the estimates obtained with the data and methods described above, we used an alternative source of data and a different estimation method to obtain age-specific ISR and ICR. Specifically, we combined age-specific reports of IFRs from the literature with the hospital and ICU mortality data listed in **Table M3** to obtain the ISR and ICR using a ratio-of-ratios method, as explained below.

Let *IFR*_*sa*_ be the expected ratio between deaths and infections estimated in a study *s* (s=1, …, S) for age stratum *a* (*a=*1, …, *A*_*s*_) and let *SFR*_*a*_ be the expected ratio between deaths and severe COVID-19 cases for age stratum *a* (*a*=1, …, *A*_*s*_). Then, we have that the estimated ISR for age stratum *a* estimated from study *s* is 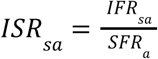. Thus, by estimating the values of *SFR* for different ages, we can use age-specific IFR values reported in the literature to obtain estimates of age-specific ISR.

To approximate the age-specific SFR, we fitted a Bayesian logistic regression to age-stratified hospital death for COVID-19 patients. Let {*d*_*la*_, *h*_*la*_} represent the number of deaths (*d*_*la*_) among *h*_*la*_ individuals hospitalized with COVID-19 for the age stratum *a* (a=1, …, A_*l*_) in location *l*. The hospital mortality for this location-stratum is defined as 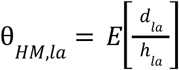.

To estimate θ_*HM*_(*age*), we used Bayesian random-effects logistic regressions, like the one described in section M4, to the hospital death data. The only difference with the procedure in M4 is that, in this case, the numerators *h*_*la*_ were known, and thus we directly used these fixed *h*_*la*_ values (unlike the *x*_*la*_ from M4, for which a distribution over possible values was obtained using seroprevalence estimates).

We use θ_*HM*_ (*a*) as our estimate of *SFR*_*a*_. These two quantities are equal if we assume that all deaths occur in the hospital (note that our definition of severe case, stated in the main text, is a case that results in either hospital admission or out-of-hospital death). As discussed in **Supplementary S3** and shown in **Figures S1, S2**, out-of-hospital deaths make only a very small fraction of severe cases for all but the oldest age-strata. Also, we find that out-of-hospital deaths make up a minority of the deaths for all but the oldest ages (analysis not shown).

Then, to account for the uncertainty of the *IFR*_*sa*_ estimates in our estimations, we fitted a Beta distribution to the mean and credible interval of each *IFR*_*sa*_ through moment matching, to obtain *IFR*_*sa*_ ∼*Beta*(α_1,*sa*_, α_2,*sa*_) (for Brazeau et. al. (2020) we only used the point estimates since credible intervals on the mean estimates are not reported).

Finally, we estimated 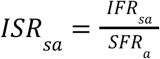 by generating samples from the posterior distribution of each *SFR*_*a*_ (obtained from the Bayesian logistic regression model) and from the Beta distribution fitted for each *IFR*_*sa*_. In total, we generated 50.000 samples of this ratio for each *ISR*_*sa*_.

The same procedure was applied to estimate the *ICR*_*sa*_, by fitting the model to ICU death data. The estimated hospital and ICU mortality rates obtained from these models are shown in **Table S3**, and the parameters obtained from fitting the model are shown in **Table S4**.

### M7) Correction for out-of-hospital and out-of-ICU deaths

Some COVID-19 deaths occur outside of the ICU, or outside of the hospital. This happens when the patient prognosis is poor, such as in elderly and frail patients, and it may be accentuated when health systems are operating at high occupancy. This phenomenon is particularly notable in our data for some locations and ages, where the number of reported deaths is larger than the number of reported ICU admissions (in some cases by more than one order of magnitude).

Our definitions of severe and critical COVID-19 outcomes include these out-of-hospital and out-of-ICU deaths, besides hospitalizations and ICU admissions. Therefore, we obtained the number of severe cases by adding to hospitalizations the number of out-of-hospital deaths. Likewise, we obtained the number of critical cases by adding to the number of ICU patients the number of out-of-ICU deaths. For some locations, we could obtain data on the out-of-hospital and out-of-ICU deaths, but for other locations this data was absent, and so we estimated it using the death data.

Let *y*_*la*_ be the cumulative number of hospitalizations for a location *l* and age stratum *a*, for which no out-of-hospital death data is available. Also, let 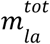 be the total number of deaths reported for this location and age stratum. First, we obtained the expected number of in-hospital-deaths, 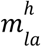, by combining the number of hospitalizations with the expected hospital mortality for this age, θ_*HM*_(*a*) (fitted as described in section M6), 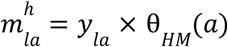. Then, we obtain the expected number of out-of-hospital 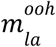 deaths by subtracting from the total number of deaths, 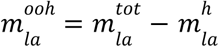 (setting 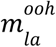 to 0 if the result is negative).

The same procedure is performed for the ICU data to obtain the number of critical cases.

See sections **Supplementary S3** and **Figures S1, S2** for an analysis showing the effect of this correction method on the data, and the robustness of the results to the removal of this correction.

### M8) Data and code availability

All data and code used in this project are available at https://github.com/dherrera1911/estimate_covid_severity.git

## Supporting information

supplementary_results

## Data Availability

All data and code used in this project are available

https://github.com/dherrera1911/estimate_covid_severity.git

## Acknowledgements

We thank Elías Eyþórsson, Gideon Meyerowitz-Katz, Nicholas Davies and Nana Owusu-Boaitey for useful discussions and comments on this work.

