## supplementary_results for "Age-specific rate of severe and critical SARS-CoV-2 infections estimated with multi-country seroprevalence studies"

### S1) Point estimation of severity for the 0-9 age range

Although the relation between disease severity and age is exponential for most ages, some evidence suggests that for the youngest ages the rates of severe disease may deviate upwards of this trend. For example, O'Driscoll et al. (2020) estimate that the IFR for ages <10 years old to be higher than expected from the exponential relation. A similar trend can be observed in the age-stratified rates of hospital admissions per 100.000 people from England (*Coronavirus (COVID-19) Latest Insights - Office for National Statistics*, n.d.). It is also apparent in our own data (**Figure 1**), where estimates from individual locations tend to fall upwards of the trend line for ages <10 years old. This is also reproduced by the estimates obtained with the indirect method, all of which fall upwards of our trend line for the youngest age brackets (**Figure 2**). Thus, given the potential of our logistic fit to underestimate the severity rates for the youngest age bracket, we estimate these rates using only data from the 0-9 age bracket for reference, in which we do not assume any relation with age.

For this, we used a Bayesian model like the one described in methods section **M4**, but removing the age effect and using only data from the single age bracket of 0-9 years old. Thus, this model only contained an intercept, with a random effect for location. After dropping the locations without data for the 0-9 age bracket, 6 locations were used to estimate the ISR (Iceland, Netherlands, New Zealand, Ontario, South Korea, Spain), 6 locations for the ICR (Netherlands, New Zealand, Ontario, South Korea, Spain, Sweden), and 7 locations for the IFR (Iceland, Netherlands, New Zealand, Ontario, South Korea, Spain, Sweden).

From this analysis, we a ISR for the 0-9 age bracket of 0.42% (95% credibility interval: [0.13-1.1]), a ICR of 0.024% [0.0039-0.076], and an IFR of 0.0014% [0.00013-0.0052], which are higher than the corresponding values shown in **Table 1**. We note that, compared to the values fitted using a regression on age, this point estimate of IFR is more similar to the estimate reported by O'Driscoll et al for the 5-9 age bracket (0.001% [0.000-0.001]) and by Levin et al for the 0-9 age bracket (0.001% [0.0007-0.0013]). Similarly, the point estimate of ISR is more in line with those obtained through the ratio-of-ratios method from O'Driscoll et al (0.25% [0.080-0.61] for the 5-9 age bracket) and from Levin et al (0.30% [0.10-0.70]). The point estimate for ICR is also closer to those obtained from the ratio-of-ratios method from O'Driscoll et al (0.029% [0.011-0.064]) and from Levin et al (0.033% [0.014-0.067]).

Although it is unclear which estimates for the 0-9 bracket are more accurate, those obtained from combining data from all ages, or those using only data from this age bracket, the close agreement between the estimates of this supplementary section and the estimates obtained from the ratio-of-ratios method supports the idea that the figures in **Table 1** may be underestimates for the younger ages.

**S2) Predicted rates**

To aid the use of the estimates obtained in this work,  we show in **Table S1** the estimates for the rates of severe, critical and fatal disease for different ages, estimated with the same method as in **Table 1** but with a finer stratification. These are the same estimates indicated by the black lines in **Figure 1** in the main text. Also, we show in **Table S2** the parameters obtained in the models used to generate **Table S1**.

Then, in **Table S3** we show the estimated age-specific values of mortality among hospital and ICU patients with COVID-19, shown in **Figure 2A**, and in **Table S4** we show the parameters for these models.

**Table S1**. Estimated values of the percentage of infected individuals that develop severe disease (ISR), critical disease (ICR) and fatal disease (IFR) for different ages. Same as **Table 1** in the main text, but with finer age stratification. (*) Note that because of the trend assumed in the fitting procedure, these estimates may underestimate the true outcome rates. See supplementary section **S1** for further discussion.

| Age | ISR % (CrI) | ICR % (CrI) | IFR % (CrI) |
|---|---|---|---|
| 0-4 | 0.086 (0.052-0.13) (*) | 0.0069 (0.0042-0.0109) (*) | 0.00036 (0.00018-0.00062) (*) |
| 5-9 | 0.13 (0.08-0.20) (*) | 0.011 (0.007-0.018) (*) | 0.00070 (0.00035-0.00121) (*) |
| 10-14 | 0.18 (0.11-0.29) | 0.019 (0.011-0.029) | 0.0014 (0.0007-0.0024) |
| 15-19 | 0.27 (0.16-0.42) | 0.030 (0.018-0.047) | 0.0026 (0.0014-0.0046) |
| 20-24 | 0.39 (0.23-0.61) | 0.050 (0.030-0.078) | 0.0051 (0.0027-0.0090) |
| 25-29 | 0.56 (0.33-0.89) | 0.081 (0.049-0.129) | 0.010 (0.005-0.018) |
| 30-34 | 0.82 (0.48-1.33) | 0.13 (0.08-0.22) | 0.020 (0.010-0.035) |
| 35-39 | 1.2 (0.7-2.0) | 0.22 (0.13-0.36) | 0.038 (0.019-0.069) |
| 40-44 | 1.7 (1.0-2.9) | 0.36 (0.20-0.60) | 0.075 (0.036-0.138) |
| 45-49 | 2.5 (1.4-4.2) | 0.59 (0.32-0.99) | 0.15 (0.07-0.27) |
| 50-54 | 3.7 (2.0-6.2) | 0.96 (0.51-1.66) | 0.29 (0.13-0.55) |
| 55-59 | 5.2 (2.8-8.9) | 1.6 (0.8-2.8) | 0.56 (0.25-1.09) |
| 60-64 | 7.5 (3.9-12.8) | 2.6 (1.3-4.6) | 1.1 (0.5-2.2) |
| 65-69 | 10.5 (5.5-18.0) | 4.1 (2.0-7.6) | 2.1 (0.9-4.3) |
| 70-74 | 14.6 (7.6-24.9) | 6.6 (3.1-12.3) | 4.0 (1.7-8.2) |

| | | | |
|---|---|---|---|
| 75-79 | 20.0 (10.4-33.2) | 10.4 (4.9-19.2) | 7.6 (3.1-15.4) |
| 80-84 | 26.6 (14.2-42.6) | 15.9 (7.5-29.0) | 13.7 (5.6-26.8) |
| 85+ | 34.3 (18.9-52.8) | 23.4 (11.3-40.9) | 23.2 (10.0-42.7) |

**Table S2**. Estimated parameters for the Bayesian logistic model with random effects describing the age-specific ISR, ICR, and IFR at different locations. The structure of the model is described in methods section M4. These parameters correspond to the fits shown in **Figure 1** in the main text.

| Model | Slope (CrI) | Slope sigma (CrI) | Intercept (CrI) | Intercept sigma (CrI) |
|---|---|---|---|---|
| ISR | 0.076 (0.068, 0.083) | 0.014 (0.009, 0.022) | -7.28 (-7.75, -6.80) | 0.86 (0.57, 1.27) |
| ICR | 0.099 (0.089, 0.108) | 0.013 (0.007, 0.023) | -9.86 (-10.34, -9.37) | 0.66 (0.38, 1.10) |
| IFR | 0.13 (0.12, 0.14) | 0.017 (0.011, 0.027) | -12.9 (-13.6, -12.3) | 1.11 (0.76, 1.59) |

**Table S3**. Estimated values of the percentage of individuals hospitalized or admitted to critical care with COVID-19 that die, for different age strata. The estimates are obtained from the fit to the hospital and ICU mortality data of **Figure 2A** in the main text. Numbers in the parenthesis indicate credible intervals of the estimates, obtained by taking the 2.5% and 97.5% quantiles of the posterior probability of the bayesian fit.

| Age | Hospital mortality % (CrI) | ICU mortality % (CrI) |
|---|---|---|
| 0-4 | 0.37 (0.13-0.83) | 3.3 (1.4-6.1) |
| 5-9 | 0.52 (0.19-1.12) | 4.1 (1.8-7.4) |
| 10-14 | 0.72 (0.29-1.50) | 5.1 (2.4-9.0) |
| 15-19 | 1.1 (0.4-2.0) | 6.3 (3.1-11) |
| 20-24 | 1.4 (0.6-2.7) | 7.8 (3.9-13.2) |
| 25-29 | 2.0 (0.9-3.7) | 9.6 (5.1-16) |
| 30-34 | 2.8 (1.4-4.9) | 11.7 (6.3-19.1) |
| 35-39 | 3.8 (2.0-6.6) | 14.3 (7.9-22.8) |
| 40-44 | 5.3 (2.9-9.0) | 17.4 (9.7-27.1) |
| 45-49 | 7.3 (4.0-12) | 20.9 (12.0-31.8) |
| 50-54 | 10.1 (5.7-16) | 25.0 (14.7-37.1) |
| 55-59 | 13.7 (7.8-21.6) | 29.5 (17.8-42.9) |
| 60-64 | 18.3 (10.6-28.4) | 34.5 (21.4-49.1) |
| 65-69 | 24.0 (14.1-36.5) | 39.8 (25.0-55.4) |
| 70-74 | 30.8 (18.5-45.6) | 45.4 (29.6-61.7) |
| 75-79 | 38.5 (23.8-55.3) | 51.1 (34.3-67.5) |
| 80-84 | 46.7 (29.8-64.6) | 56.7 (39.3-72.9) |
| 85+ | 55.1 (36.6-73.0) | 62.1 (44.5-77.9) |

**Table S4**. Estimated parameters for the Bayesian logistic model with random effects describing the age-specific hospital and ICU mortality. The structure of the model is described in methods section M5. These parameters correspond to the fits shown in **Figure 2A** in the main text.

| **Model** | **Slope (CrI)** | **Slope sigma (CrI)** | **Intercept (CrI)** | **Intercept sigma (CrI)** |
|---|---|---|---|---|
| Hospital mortality | 0.068 (0.054, 0.082) | 0.017 (0.009, 0.033) | -5.8 (-6.7, -4.9) | 1.2 (0.7, 1.9) |
| ICU mortality | 0.045 (0.035, 0.056) | 0.0097 (0.0028, 0.0246) | -3.5 (-4.2, -2.9) | 0.82 (0.44, 1.43) |

#### S3) The correction method for out-of-hospital and out-of-ICU deaths is in agreement with real-world data

As described in methods section **M7**, we applied a correction to some of the data points, where we estimated the out-of-hospital and out-of-ICU deaths in order to obtain the total severe and critical cases. In **Figure S1** we show the data with and without this correction (left and right panels, respectively), and the estimated rates obtained by fitting the model to each dataset. Also, the locations for which actual data on out-of-hospital and out-of-ICU deaths was available, and thus did not need to be corrected with estimated out-of-hospital and out-of-ICU deaths, are indicated with dashed lines and triangles.

Comparing the plots from the top row, which correspond to the corrected and uncorrected rates of severe disease, we observe that the correction procedure has very little effect on the severe infection rates overall, with only minor changes for the oldest ages. The fitted model also remained mostly unchanged.

Comparing the plots of corrected and uncorrected rates of critical disease (bottom row), we observe that the correction procedure has a large effect for older ages, but only a small effect for younger ages (i.e. for age bins under 50 years). This is in line with the expectations that out-of-ICU deaths are rare for younger ages, but common for older ages. Furthermore, we see that the different locations cluster into two groups in the plot of the uncorrected critical disease rate (lower right). For countries where out-of-ICU death data was available (and used to obtain the critical cases), the rate of critical disease follows a log-linear trend, while the countries without this data (and thus with only ICU admissions used on the bottom right plot) show a drop in the critical rate. Thus, the observation that using estimated out-of-ICU deaths for countries without available data aligns them with the countries where this data is available, suggests that the correction for out-of-ICU deaths correctly captures the numbers of out-of-ICU deaths.

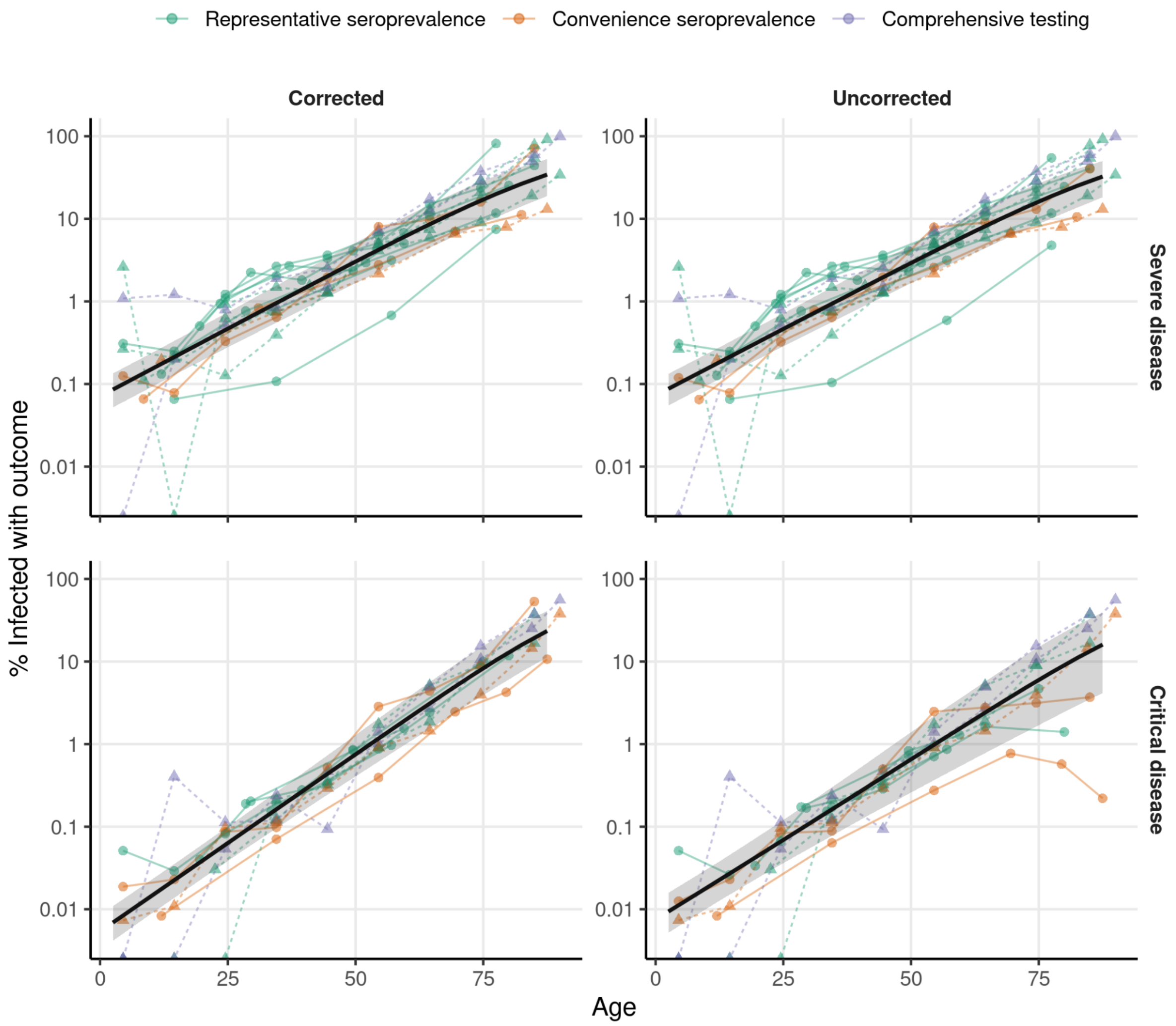

**Figure S1**. **Correcting for out-of-hospital and out-of-ICU deaths has a small effect on the overall fit.** Data for the ISR (top) and ICR (bottom), with (left) and without (right) correcting for out-of-hospital and out-of-ICU deaths are shown. Each dot shows the ISR or ICR for one location and age, and the different ages for one given location are connected by the lines. The color of the lines and dots indicates the method used to estimate the number of infected individuals. The black lines show the fitted logistic regression (in logarithmic scale, as in **Figure 1** of the main text). The data shown in triangles and joined with dashed lines corresponds to locations to which correction for out-of-hospital or out-of-ICU deaths was applied using real-world data for these locations, instead of the estimation method. In these cases, the numbers of severe or critical disease are the same for the corrected and uncorrected datasets.

To further evaluate the precision of the correction method, we compared the proportion of severe and critical cases that consist of out-of-hospital and out-of-ICU deaths for countries with real-world data, and for countries where these deaths were estimated. If the proportions obtained with the estimated quantities match the proportions obtained from real-world data, that would suggest that our estimation method is accurate.

In **Figure S2A** we see that, as expected, the percentage of severe cases comprised by out-of-hospital deaths is small for young and middle-aged populations. For older populations we see considerable variability between locations, but that the percentages obtained from real data (red) and the percentages estimated with the correction method (blue) are within the same range of values.

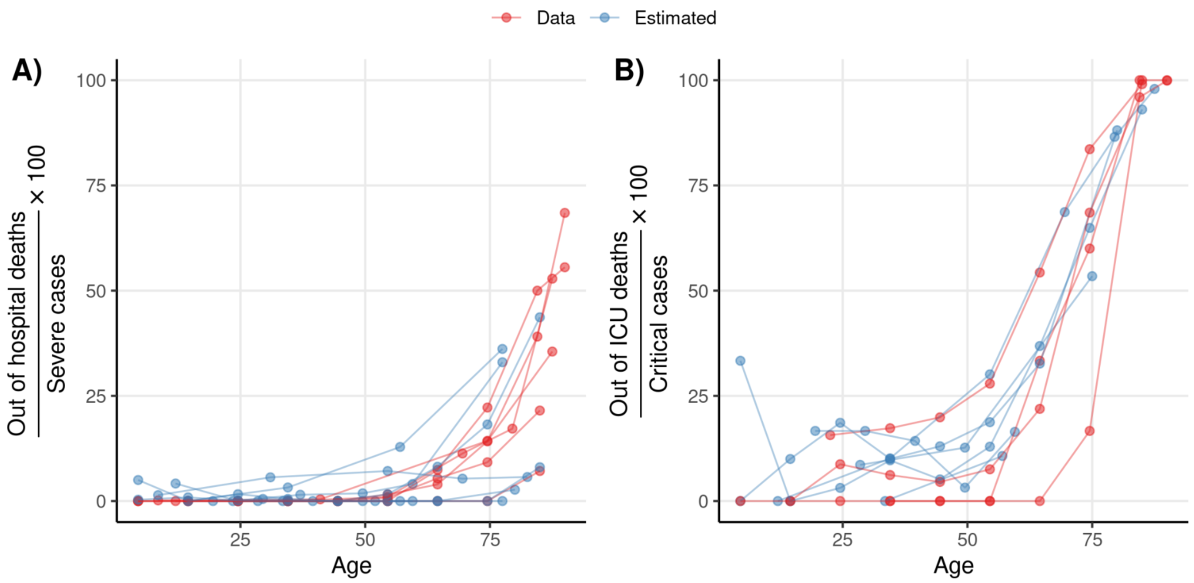

**Figure S2. Real and estimated data agree on the proportion of out-of-hospital and out-of-ICU deaths among severe and critical cases. A)** The percentage of severe cases that correspond to out-of-hospital deaths are shown for all locations with either available data (shown in red), or where these deaths were estimated with the correction procedure detailed in the methods (shown in blue). The percentages of the different ages from a given location are connected by lines. **B)** Same as **A)** but for the percentage of critical cases that are out-of-ICU deaths.

Likewise, **Figure S2B** reveals that the proportion of critical cases comprised by out-of-ICU deaths is relatively small for young ages, but that these rapidly increase for older ages, becoming more than half of the critical cases at around 65 years old, and almost all critical cases over 75 years old. While there is variability between locations, overall, there is a clear agreement in trend between locations, including locations for which the percentages were estimated from real-world data (red), and the locations with the percentages were estimated using the correction method (blue). We note that for some locations, the real-world data of hospital or ICU deaths and the data on total deaths, which were put together to estimate out-of-hospital and out-of-ICU deaths, were obtained from different sources. Therefore, some of the out-of-hospital or out-of-ICU deaths estimated from real-world data may also reflect differences in reporting delays and methods between the sources.

In conclusion, we observe that the correction method induces only small changes to severe cases for all but the oldest ages and that it induces relatively small changes to critical cases at young ages, but that out-of-ICU deaths increase steeply at older ages until they comprise most of the critical cases. Also, we observe that the correction from our estimation method matches what is seen in the locations where data on out-of-hospital and out-of-ICU deaths is available.

**S4) Regression results are robust to excluding older ages**

Next, given that we argue for the usefulness of our estimates of ISR and ICR for the cost-benefit analysis of children and adolescent vaccination, we tested whether our estimates for these ages are robust to excluding the data from the oldest ages. If our estimates for younger ages are not affected by excluding older ages, this would suggest that the data at younger ages alone is sufficient to accurately estimate these outcome rates.

In **Figure S3** we show in black the same model as the one in **Figure 1** in the main text. Then, overlapped with the main model, we show in red and in blue the models fitted to the uncorrected data (same as the right panels in **Figure S1**), but excluding age bins with a median age over 40 (red) and over 50 (blue) respectively. We see, the estimates for ISR (left), ICR (center), and IFR (right), change little when fitting the models to the different subsets of the data, and that in all cases the estimates have mostly overlapping credible intervals. This indicates that our estimates are robust to different preprocessing choices, and to using only younger ages for the estimation. Therefore, we conclude from this analysis that our estimations of ISR, ICR, and IFR for younger ages are not significantly affected by the data from older ages and that they are robust to changes in the preprocessing of the data.

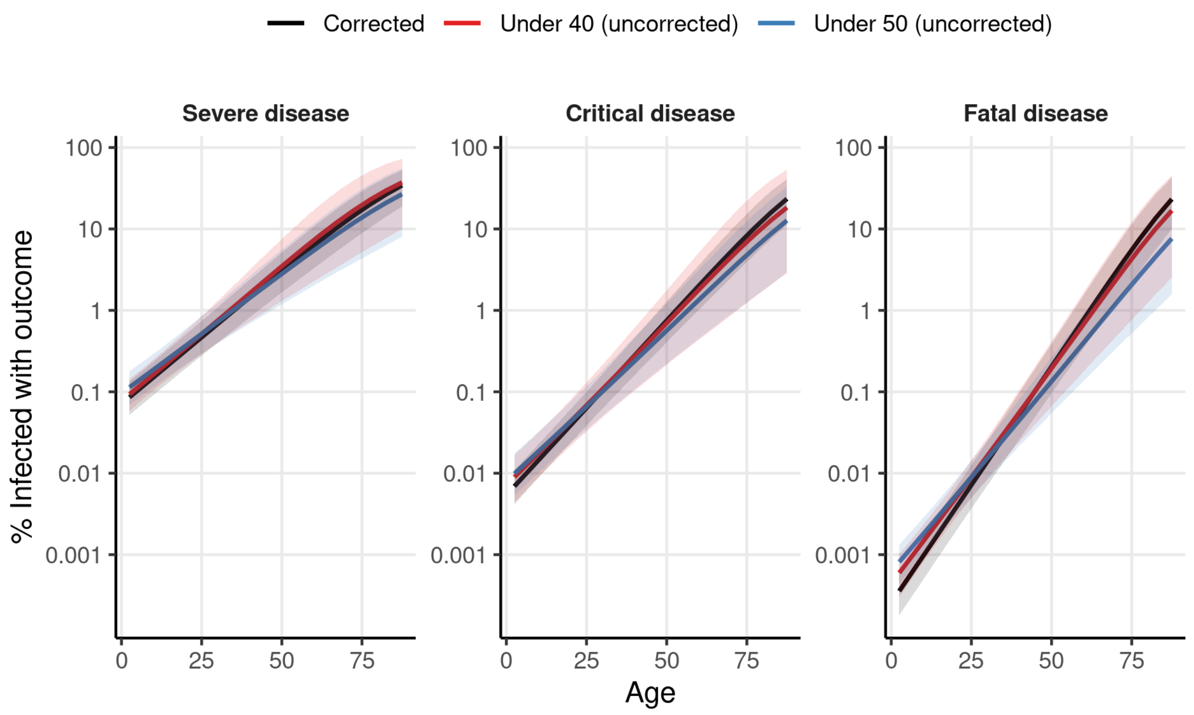

**Figure S3. ISR, ICR, and IFR estimations are robust to excluding older ages.** Lines show the estimated ISR (left), ICR (center), and IFR (right) obtained from fitting the Bayesian logistic regression model (as in **Figure 1** in the main text) after different data preprocessing procedures. The black line shows the main model, fitted to the same data as **Figure 1** in the main text. The red and blue lines show the models fitted to the uncorrected data (same as right panels of **Figure S1**), but only using the data from age bins with a median age under 40 and under 50 years, respectively. Colored shaded regions show the 95% credible interval for each estimate.

### S5) Regression results are robust to excluding data from comprehensive testing or from convenience seroprevalence

Also, our analysis includes locations with three different methods of estimation of SARS-CoV-2 prevalence: serosurveys with population-representative samples, serosurveys with convenience samples, and comprehensive testing and tracing. Of these, the serosurveys with representative samples are expected to be the most accurate, while the other two methods can possibly introduce a bias into the prevalence estimates. Therefore, we next tested whether our results are robust to excluding data from locations with convenience serosurveys, and from the locations with comprehensive testing.

We see in **Figure S4** that the estimates of the ISR (left), ICR (center) and IFR (right) obtained from the different subsets of the data are in close agreement with the estimates obtained from the full dataset. Thus, we conclude that our results are not biased by the studies from which SARS-CoV-2 infections were estimated from convenience seroprevalence studies, or from comprehensive testing data.

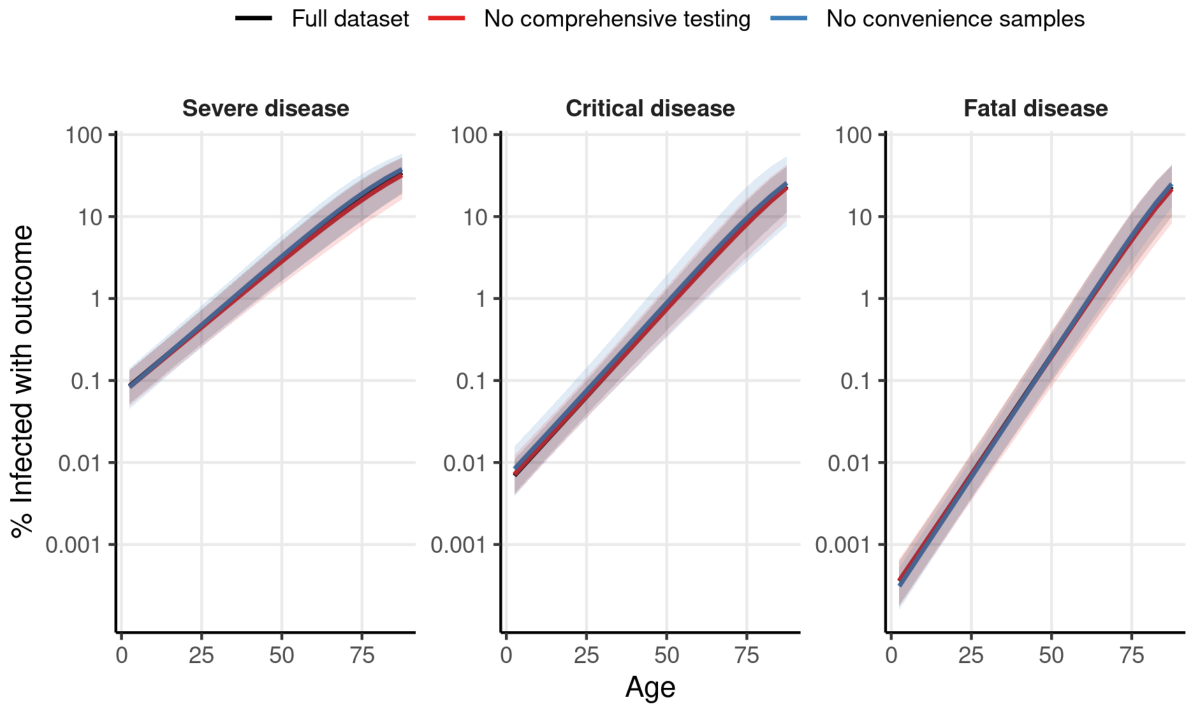

**Figure S4. ISR and ICR estimations are robust to excluding data from serosurveys with convenience samples and comprehensive testing.** Lines show the estimated ISR (left), ICR (center), and IFR (right) obtained from fitting the Bayesian logistic regression model (as in **Figure 1** in the main text) to different subsets of the data. The black line shows the main model, fitted to the same data as **Figure 1** in the main text. The red line shows the model fitted to the dataset excluding locations with prevalence estimated from comprehensive testing. The blue line shows the model fitted to the dataset excluding convenience sample serosurveys.

#### S6) Epidemic dynamics at near the date of data collection

One important factor that needs to be taken into account for estimating the rate of a given outcome among infected individuals is that there is a delay between the infection and the outcome. Thus, the cumulative infections up to a certain date should be matched with the number of outcomes at a later date, to account for this delay, and this is particularly important for growing epidemics.

However, other delays also affect the data of our analysis in ways that make it difficult to precisely identify at which dates the reported disease outcomes match the infections estimated in the serosurveys. For example, there is a delay between infection and the generation of detectable antibodies (seroconversion), making serosurveys reflect the infections at an earlier date than when they are conducted. This is further complicated by the fact that the collection of samples in each serosurvey is done over a certain period, making it unclear what date to take as a reference for the serosurvey. Furthermore, there are delays between the dates at which disease outcomes occur and the dates at which they are reported in official figures. These factors also change from location to location (and even between data sources within locations), making it challenging to choose a single delay period to use between the dates of serosurveys and the dates at which we collect disease outcomes data.

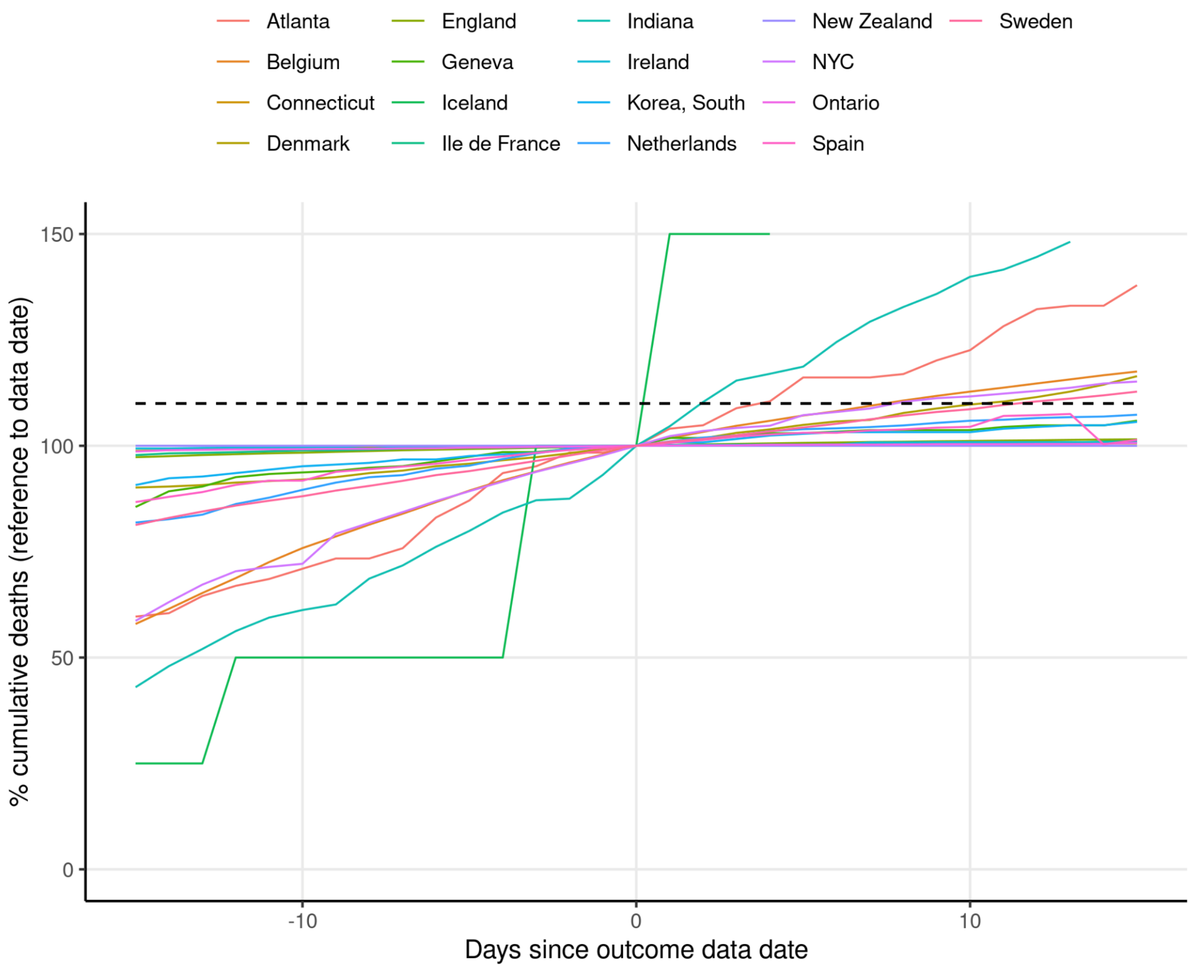

**Figure S5. COVID-19 death dynamics around the date of outcome data collection.** In this plot, each colored line shows the number of cumulative COVID-19 deaths from 15 days before the date at which outcome data was collected for the main analysis, to 15 days after this date, relative to the number of cumulative deaths at the date of data collection (indicated by day 0 in the horizontal axis). Each colored line shows the deaths for a different location used in the analysis. The horizontal dashed line indicates the value of 110%.

To collect the outcome data, we chose the simple procedure of collecting the data for disease outcomes at the date at which sample collection for the seroprevalence studies was finished (or, for locations with comprehensive testing, the date of the reported number of diagnosed cases). Because of the reasons discussed above, however, this may produce some bias in our estimates in unpredictable ways. However, the delays mentioned above are expected to be a problem only in more rapidly changing epidemics. After an epidemic has peaked and declined, the change in the number of cumulative outcome numbers will be small, and thus it will make little difference what precise date the outcomes are collected at.

Although the list of seroprevalence studies used in our analysis was already curated by Levin et al (2020) to exclude studies performed during rapidly changing epidemics, we further analyzed whether our estimates may be affected by these factors. For this, we first plotted the relative change in the number of cumulative deaths around the date of data collection (Guidotti & Ardia, 2020), shown in **Figure S5**. We can see in **Figure S5** that the deaths at the locations used had relatively small changes around the dates where we collected the data, suggesting that the date of data collection should have a small impact on the results (not that the large change seen for Iceland in the relative number of cases represents only a few absolute cases). However, we see that 4 locations (Atlanta, USA; New York City, USA; Indiana, USA; Belgium; Sweden; Denmark) have an increase in the relative number of deaths of 10% within 15 days of the outcome data report date. Therefore, to verify that these more rapidly changing locations are not biasing our estimated outcome rates, we fitted the models again excluding data from these 6 locations.

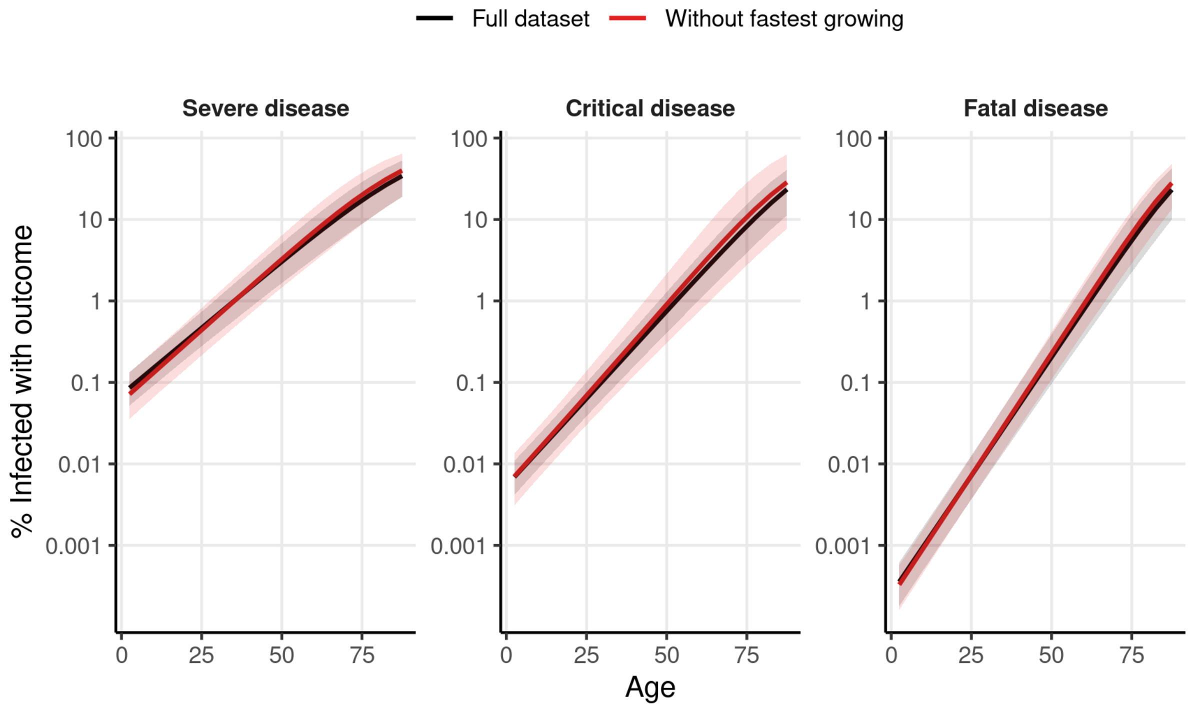

**Figure S6. ISR, ICR, and IFR estimates are robust to excluding fastest changing epidemics.** Lines show the estimated ISR (left), ICR (center), and IFR (right) obtained from fitting the Bayesian logistic regression model (as in **Figure 1** in the main text) after different data preprocessing procedures. The black line shows the main model, fitted to the same data as **Figure 1** in the main text. The red line shows the model fitted to the dataset excluding data from the locations where the cumulative death count increases 10% or more within 15 days of our outcome data collection date.

We see in **Figure S6** that the estimates obtained with the model change little when excluding the 6 locations with the fastest growing epidemics, and that the credible intervals of the two fits have a large overlap. Since we expect any biases introduced by the dates of outcome data collection to be mostly mediated by the regions with the fastest growing epidemics, we conclude that the choice of the precise date of data collection has a small effect on our results.

**S7) Effect of time between epidemic wave and seroprevalence data collection**

Another important factor that can also affect serology-based estimations of outcome rates is the phenomenon of seroreversion. Seroreversion consists of the natural decay of antibody levels with time, which can lead serological tests to miss this indicator of previous infection after a certain time. Although some studies show that IgG antibodies are developed by the majority of individuals with SARS-CoV-2 infection (Glück et al., 2021; Long et al., 2020; Wajnberg et al., 2020), and that these remain detectable in the serum for several months

(Glück et al., 2021; Iyer et al., 2020; Wajnberg et al., 2020), other studies have reported faster seroreversion, which may result in considerable percentages of seropositive individuals reverting to a seronegative result within few months of their initial antibody response (Long et al., 2020; Orth-Höller et al., 2021; Self et al., 2020). This may raise concerns about the validity of serosurveys to estimate the number of infections in a population if there is a considerable delay between the epidemic waves and the serosurvey sample collection.

To check whether our results may be biased by seroreversion, we first analyzed the delays between the epidemic waves and the serosurvey data collection across the locations. Similar to section **S6**, we analyzed the dynamics of COVID-19 deaths for the different locations. Although deaths are a lagging indicator of infections in a population, so is the presence of IgG antibodies in the population. The mean delay from symptoms onset to death is estimated to be around 15 days (Khalili et al., 2020), and the median delay between symptoms onset and maximal IgG levels has been reported to be 25 days for IgG (Iyer et al., 2020). Thus, for simplicity, we take reported deaths in a population to reflect approximately the times at which individuals in a population become maximally seropositive.

In particular, we looked at what percentage of the cumulative deaths reported at the final date of the serosurvey had occurred in the prior 45 days. Locations in which this percentage is high, indicate that the serosurvey was performed close to the date in which a large portion of the individuals had achieved maximum levels of IgG, and so seroreversion is expected to be relatively small by the date of the serosurvey, even according to studies that have reported faster estimates of seroreversion (Long et al., 2020; Orth-Höller et al., 2021; Self et al., 2020).

We find that for 10 of the locations used, at least 70% of the cumulative deaths by the date of the serosurvey had occurred in the 45 previous days (Iceland, Republic of Korea, Spain, Sweden, Netherlands, Atlanta, New York City, Geneva, Indiana and Belgium). We thus identify these locations as having a low risk of bias from seroreversion, and we fitted the Bayesian logistic regression model to these locations. We observe in **Figure S7** that the fit obtained from these locations is very similar to the fit obtained from the full dataset. This suggests that seroreversion is not a major source of bias in our estimates. This result is in line with the IFR estimates from Brazeau et al., (2020), which show little change when correcting for seroreversion.

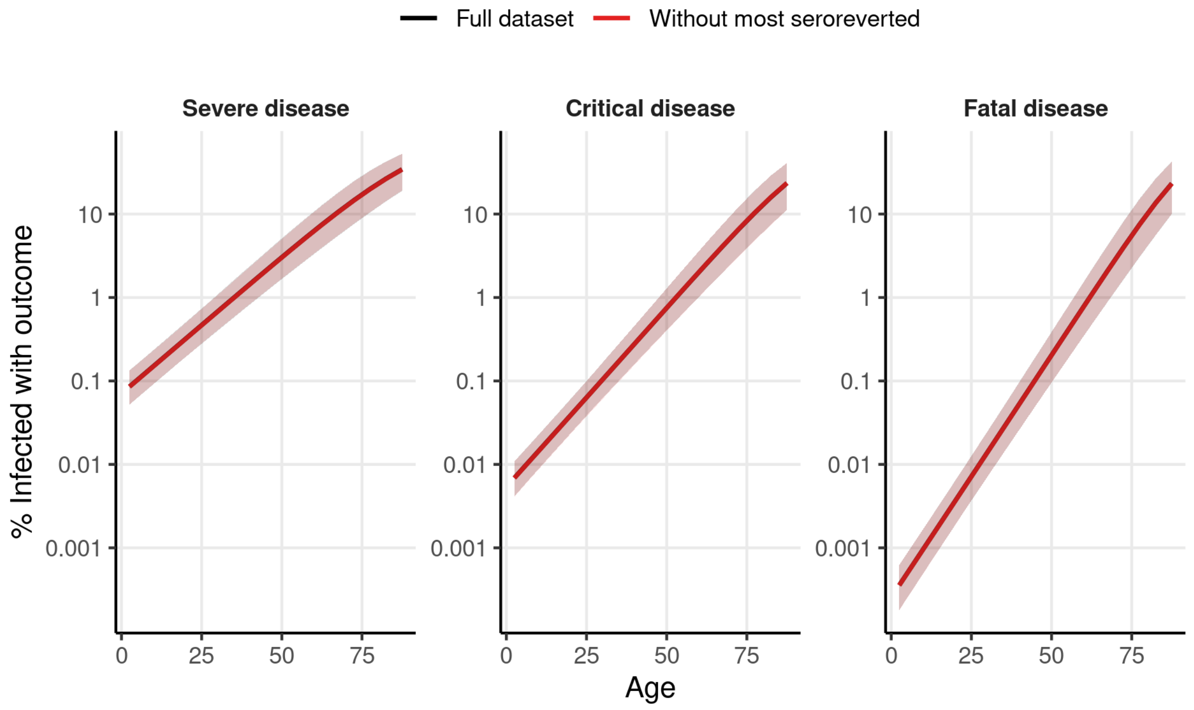

**Figure S7. ISR, ICR, and IFR estimates are robust to excluding locations with the largest epidemic to serosurvey delays.** Lines show the estimated ISR (left), ICR (center), and IFR (right) obtained from fitting the Bayesian logistic regression model fitted to seroprevalence data. The black line shows the main model, fitted to the same data as **Figure 1** in the main text. The red line shows the model fitted to the dataset including only the 10 locations in which over 70% of the cumulative deaths at the time of the serosurvey occurred within the 45 days prior to seroprevalence data collection.
